# Association of inflammatory markers with severity of disease and mortality in COVID-19 patients: a systematic review and meta-analysis

**DOI:** 10.1101/2021.09.16.21263678

**Authors:** Sara Khan, Pinki Mishra, Rizwana Parveen, Ram Bajpai, Mohd Ashif Khan, Nidhi Bharal Agarwal

## Abstract

**Purpose:** Literature suggests association of inflammatory markers with the severity and mortality related to COVID-19, but there are varying conclusions available. We aimed to provide an overview of the association of inflammatory markers with the severity and mortality of COVID-19 patients.

**Methods:** We searched Medline (via PubMed), Cochrane, Clinicaltrials.gov databases until Sept 1, 2020.

**Results:** A total of 21 studies comprising 4023 patients with COVID-19 were included in our analysis. Levels of IL-6 (WMD=18.17 95%CI 3.38 to 32.96, p=0.016), IL-8 (WMD=12.09 95%CI 4.41 to 19.77, p=0.002), MCP-1 (WMD=146.66 95%CI 88.16 to 205.16, p<0.001), CRP (WMD=31.09 95%CI 10.08 to 52.10, p=0.004), PCT (WMD= -31.23 95%CI -37.70 to -24.76, p<0.001), IL-2R (WMD=861.93 95%CI 275.45 to 1448.41, p=0.004), ferritin (WMD= 1083.34 95%CI 431.99 to 1734.70, p=0.001) were found significantly higher in the severe group compared with the non-severe group of COVID-19 patients. Moreover, non-survivors had a higher levels of IL-2R (WMD= -666.06 95%CI -782.54 to -549.59, p<0.001), IL-8 (WMD= -26.63 95%CI -33.031 to -20.236, p<0.001), IL-10 (WMD= -7.60 95%CI -8.93 to -6.26, p<0.001), TNF-α (WMD= -4.60 95%CI -5.71 to -3.48, p<0.001), IL-1β (WMD=22.66 95%CI 8.13 to 37.19, p=0.002), CRP (WMD= -96.40 95%CI -117.84 to -74.97, p<0.001), and ferritin (WMD= -937.60 95%CI -1084.15 to -791.065, p<0.001) when compared to the non-survivor group.

**Conclusion:** This meta-analysis highlights the association of inflammatory markers with the severity and mortality of COVID-19 patients. Measurement of these inflammatory markers may assist clinicians to monitor and evaluate the severity and prognosis of COVID-19 thereby reducing the mortality rate.

## INTRODUCTION

In December 2019, a largely enveloped virus was labeled as COVID-19, which was discovered in Wuhan, China, rapidly spread globally. It was found out to be a new infectious disease classified under SARS-CoV and MERS-CoV which primarily causes respiratory tract infection ^1^. WHO declared the pandemic outbreak of novel coronavirus (2019-nCoV) as a global health emergency. As of September 1, 2020 over 216 countries have been affected by the COVID -19 disease with more than Twenty Four million (24,257,989) confirmed cases leading to over 8,27,246 deaths ^2^.

Although, most cases are mild to moderate, some patients develop severe symptoms characterized by respiratory dysfunction and/or multiple organ failure ^3,4^. Identification of progression of COVID-19 in the current state, depends mostly on the clinical manifestation. It has been recommended that one of the possible mechanisms of ultimate speedy disease progression is cytokine storm ^5,6^. Cytokine storm, also known as cytokine cascade, or hypercytokinemia, is caused by infection, drugs or autoimmune diseases of the body’s excessive immunity response ^7^. This inflammatory response is associated with elevated levels of inflammatory cytokines and leads to alveolar damage and acute respiratory distress syndrome (ARDS) ^5^. Accumulating evidence suggest that an elevated level of inflammatory cytokines is associated with a high severity and case fatality of COVID-19 infection ^4,6,8–10^. Pathological manifestations of COVID-19 greatly resemble what has been seen in SARS and MERS infection where massive interstitial inflammatory infiltrates diffuse in the lung ^10^. The main effect of interleukins is to suppress inflammation, however, it has been shown that the over-production of specific inflammatory cytokines such as IL-2, IL-4, IL-6, IL-8, IL-10, tumor necrosis factor (TNF-α) are the gold standard of viral infection^11–13^. Interleukins can be of high interest for reaching out the target of immunosuppressive therapy approaches by cytokine blockers in COVID-19 like several approaches, including global targeting of the inflammation or neutralizing a single key inflammatory mediator, can be employed to cope with cytokine storm ^14^. Interleukins promotes specific differentiation of CD4^+^ / CD8^+^Tcells, thus performing an important function in the linking of innate to acquired immune response. Also, in combination with transforming growth factor (TGF)-β, interleukins are indispensable for Th17 differentiation from naıve CD4^+^ T cells^15^. Elevated levels of IL-6 can cause hyper-activation of JAK/STAT3 signaling, which is often associated with poor patient outcomes ^16^.

The elevated cytokine levels may also be responsible for the lethal complications of patients with COVID-19, SARS or MERS presented distinct cytokine profiles^17^. Along with interleukins the levels of C-reactive protein(CRP), induced by IL-6, is a significant biomarker of inflammation, infection and tissue damage ^18^ along with Procalcitonin (PCT) that is a glycoprotein and is the precursor of calcitonin, its levels are increased with bacterial and viral levels and can be indicative of the disease severity as these levels were found to be greater in severe group of patients than in mild group ^9^. Ferritin, a protein that contains iron and is the primary form of iron stored inside of cells. Levels of ferritin could be a significant marker for COVID-19 as it is found elevated in diseased patients ^19^.

To the best of our knowledge, due to the insufficient sample sizes, the complete inflammatory profile is missing to date. Therefore, we did a meta-analysis based on the current literature to compare the levels of inflammatory markers between severe vs non-severe and survivor vs non-survivors patients with COVID-19. Our findings will give insight about the role of inflammation in the pathophysiology of novel coronavirus and its clinical features. Detection of these inflammatory markers can work as a tool to group patients according to their disease severity and predict the prognosis and mortality, thereby contributing to treatment and reducing mortality rate.

## MATERIALS AND METHODS

Preferred Reporting Items for Systematic Review and Meta-Analysis (PRISMA) guidelines extension for scoping reviews^20^ and MOOSE guidelines were followed for designing and reporting this systematic review. The protocol of this systematic review was registered on PROSPERO (registration ID: CRD42020200757).

### Data sources and searches

A systematic literature search was performed using Medline (via PubMed), The Cochrane Central Register of Controlled Trials (CENTRAL) and Clinicaltrials.gov until September 1, 2020 using the keywords “SARS CoV-2”, “Interleukins”, “Cytokines”, “COVID-19”, “Cytokine storm”, “Inflammatory markers”, “laboratory findings”. We also searched grey literature using Google Scholar and reference list of eligible articles with the aim of identifying additional potential eligible studies. Each article was separately reviewed by two authors independently to find out the right content. The searches were limited to articles published in English language.

### Inclusion and exclusion criteria

Observational studies or case series reporting the level of cytokines in defined groups as per severity of COVID-19 were included. We excluded duplicate publications, reviews, editorials, case reports, letters, meta-analysis, systematic review, protocols, studies in language other than English and studies not reporting the required data. First author (SK) searched data and screened articles for eligibility. Senior author (PM) double checked all the included articles and any dispute was resolved by the third author (RP).

### Quality assessment

Two reviewers (SK and PM) assessed the quality of data in the included studies using the National Institute of Health (NIH) quality assessment tools ^21^. We preferred the NIH tool because it is comprehensive and widely accepted for an exhaustive assessment of data quality. We rated the overall quality of included studies as good, fair and poor, and incorporated them in the results of this systematic review. The tools were designed to assist reviewers in focusing on concepts that are key for critical appraisal of the internal validity of a study. The tools were not designed to provide a list of factors comprising a numeric score. The tools were specific to individual types of included study designs and are described in more detail below. The tools included items for evaluating potential flaws in study methods or implementation, including sources of bias (e.g., patient selection, performance, attrition, and detection), confounding, study power, the strength of causality in the association between interventions and outcomes, and other factors. Quality reviewers could select “yes,” “no,” or “cannot determine/not reported/not applicable” in response to each item on the tool. For each item where “no” was selected, reviewers were instructed to consider the potential risk of bias that could be introduced by that flaw in the study design or implementation. Cannot determine and not reported were also noted as representing potential flaws. Each of the quality assessment tools had a detailed guidance document, which was also developed by the methodology team and NHLBI.

### Data Extraction and analysis

Data was inputted into a standardized data extraction table (Excel) and independently checked by a second reviewer (PM) for accuracy. The following variables were extracted: name of the first author, year of publication, study design, age, gender, number of patients in severe and non-severe and survivor vs non-survivor groups, comorbidities, and the reported level of cytokines. The severity of disease was defined according to Diagnosis and Treatment Plan of COVID-19 issued by National Health Commission, China (7th edition) as mild, common, severe, and critical based on the clinical symptoms^22^. The articles that were included in this systematic review are displayed in PRISMA flow diagram **(Figure-I)**.

**Figure-I.**
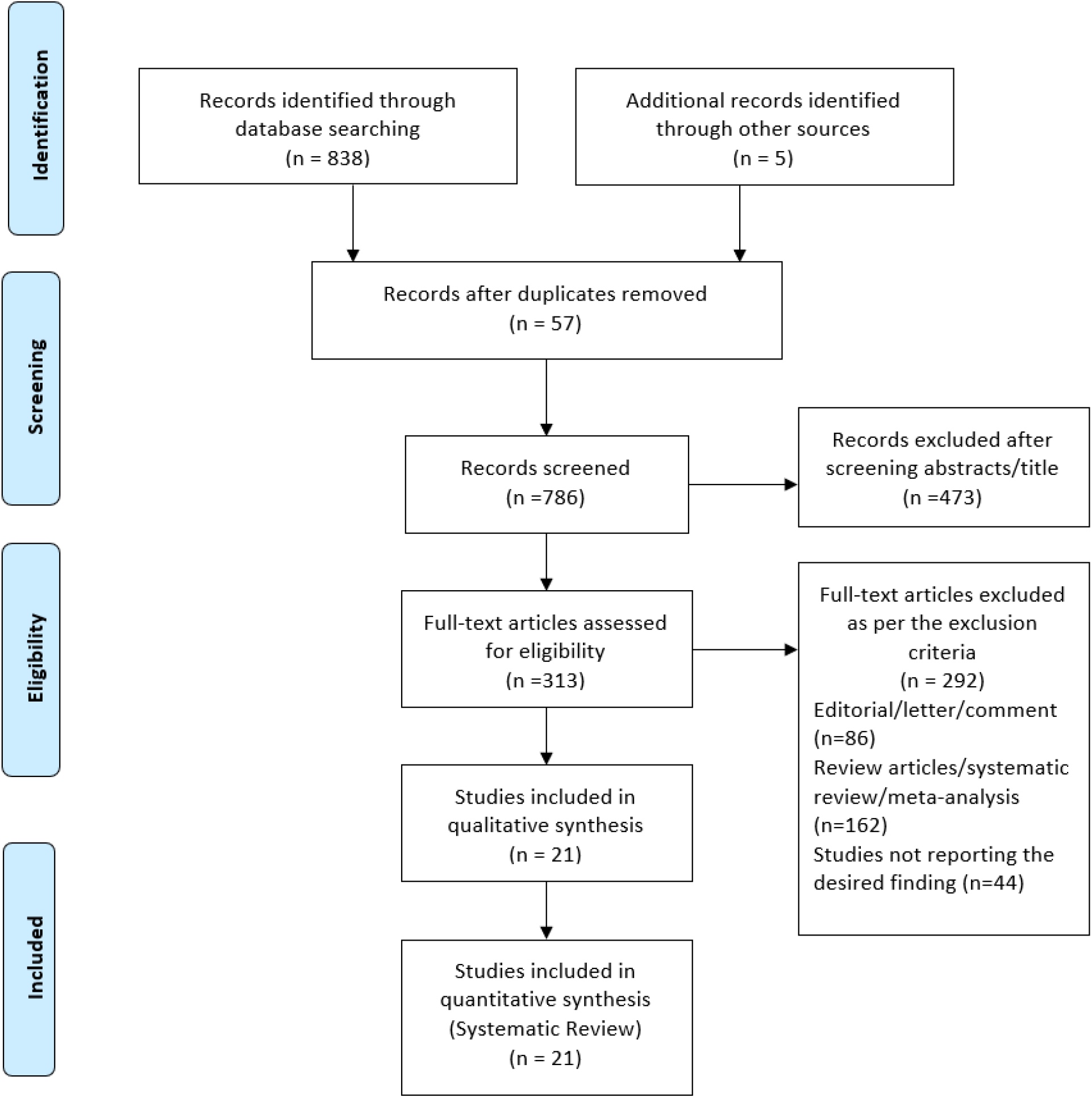
PRISMA flow diagram

### Outcomes

The primary outcomes were (1) Severity of COVID-19 including: Intensive Care Unit (ICU) admission and (2) mortality due to confirmed COVID-19. Only intra-hospital mortality was considered.

### Analysis plan

We performed an exploratory meta-analysis to understand the magnitude and direction of effect estimate. Continuous outcomes are presented using weighted mean difference (WMD) and 95% confidence intervals (CIs). Odds ratios (ORs) were calculated and presented with respective 95% CIs for binary outcomes. Meta-analysis of ORs using Mantel-Haenszel random-effects and inverse-variance method for WMDs using DerSimonian-Laird method was used ^23^. Heterogeneity between studies was assessed using the χ^2^-based Cochran’s Q statistic (p<0.1 considered as the presence of heterogeneity) and I-squared (*I*^*2*^) statistics (>50% representing moderate heterogeneity) ^23^. Publication bias was not assessed as a total number of studies for respective outcomes were less than ten ^23^.

## RESULTS

### Search Results

The systematic search yielded a total of 838 publications. After removing duplicates, 313 articles were found to be potential publications for screening. After the application of predefined inclusion and exclusion criteria, a total of 21 studies were included for the final analysis **(Figure-I)**.

### 3.1 Study characteristics

Among the 21 included studies, 16 were cohort studies, while 5 were case series. The main characteristics of the included studies are reported in **Table-I**. Overall, 4,023 participants were enrolled in the present study, out of which 2,190 (57.55%) were male and 1,615 (42.44%) were females. The patients were classified into 4 groups on the basis of their disease severity: severe and non-severe or survivor and non-survivor group. Severe group had a total of 1,242 patients (32.64 %) whereas non –severe group had 2,563 patients (67.35 %). Survivor group had 425 patients (62.9%) and Non-Survivor group had 250 patients (37.03%). Inflammatory markers that were assessed in the present systematic review were IL-2, IL-2R, IL-4, IL-6, IL-8, IL-10, IL-1β, TNF-α, MCP-1, C-reactive protein, procalcitonin, and ferritin.

**Table-I.**
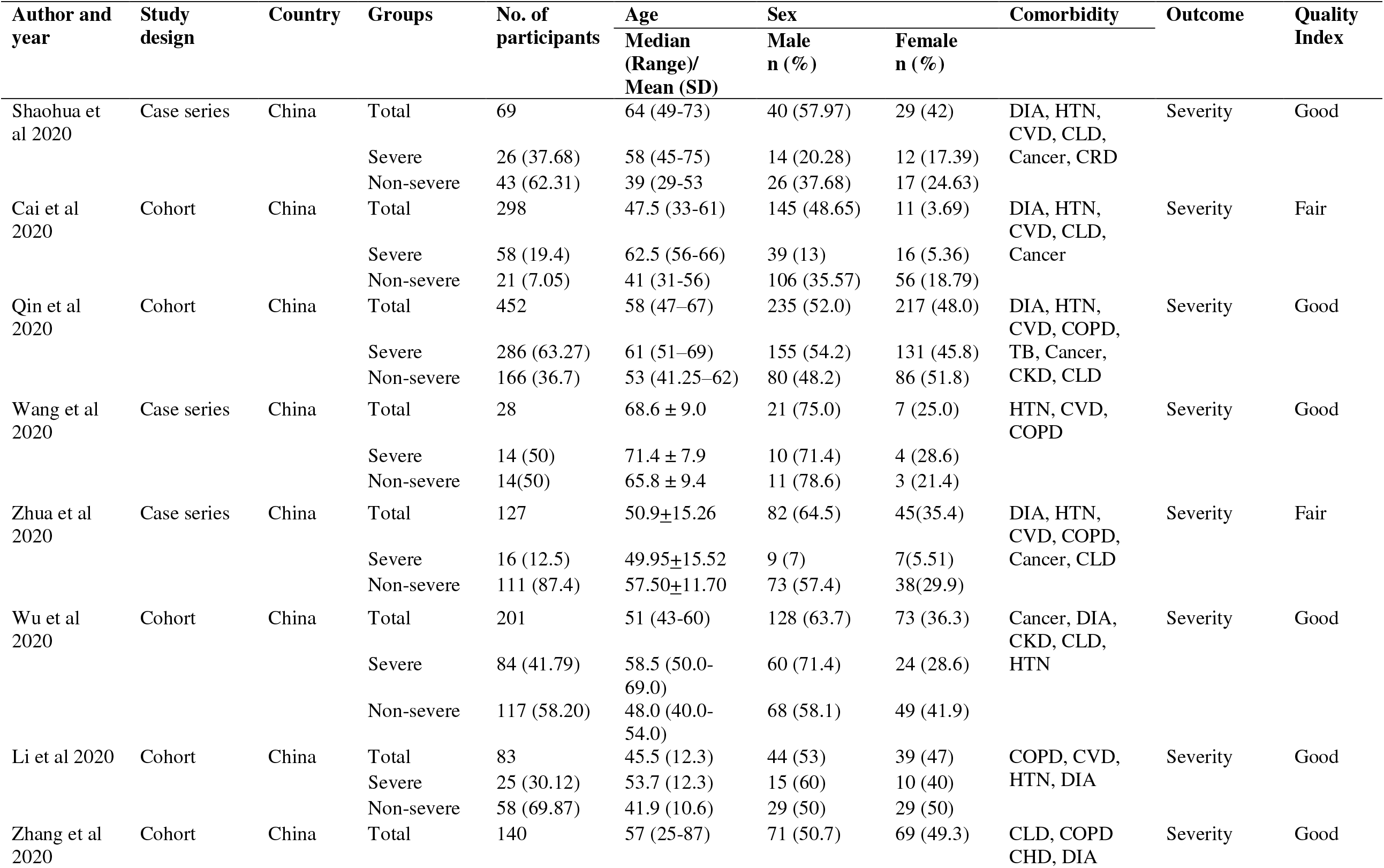
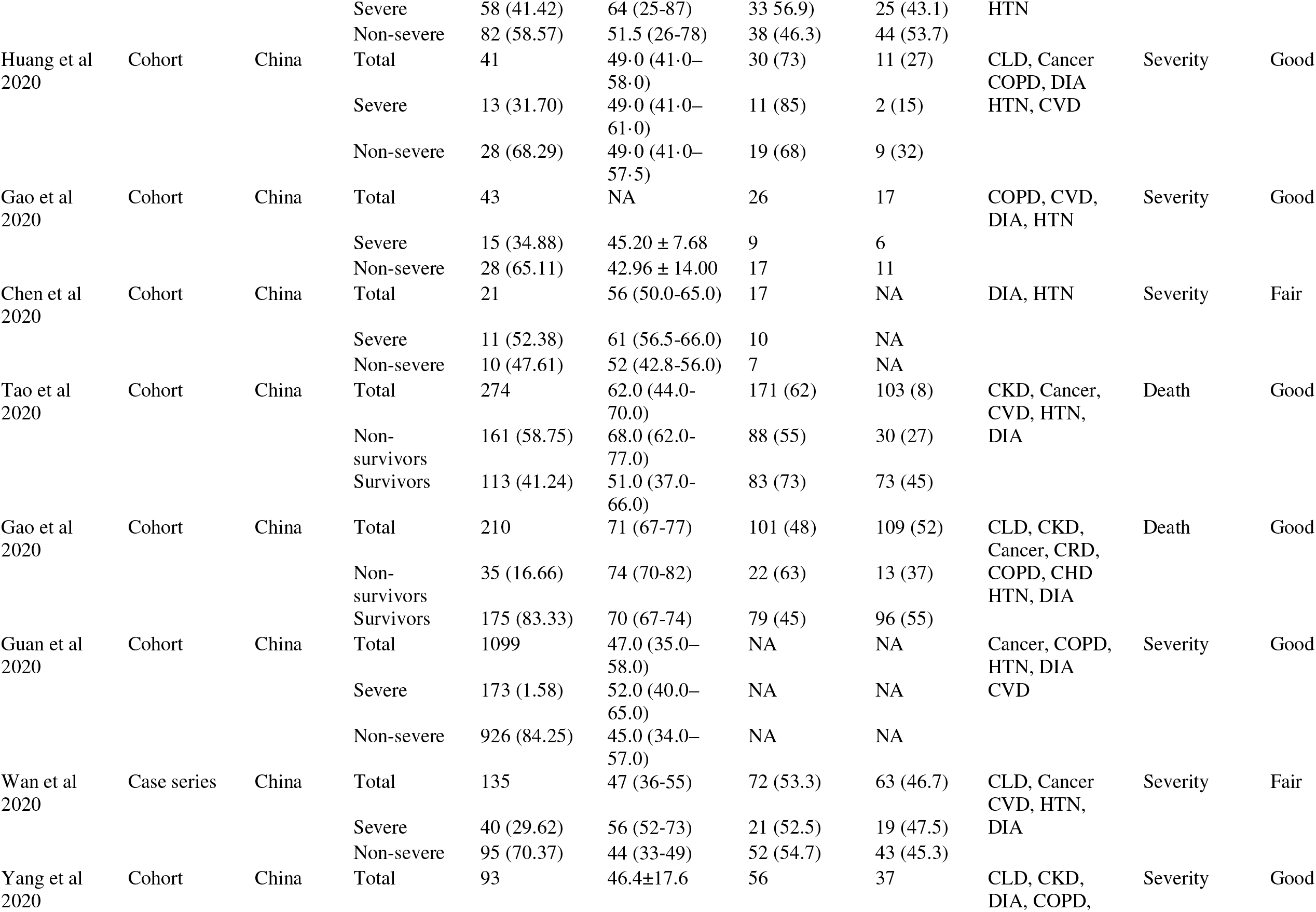
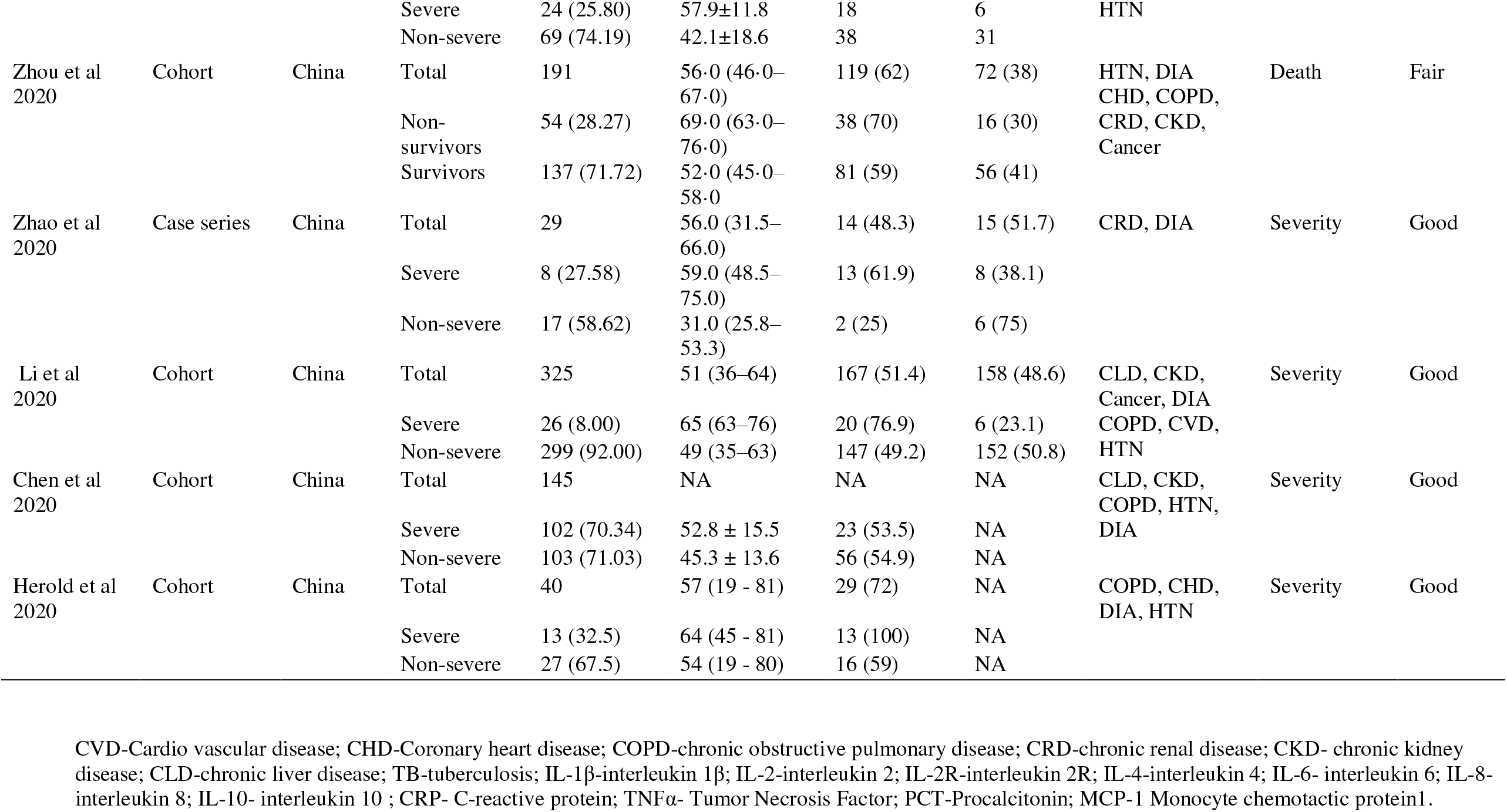
Study Characteristics.

### 3.2 Association of Cytokines with disease severity

For the patients stratified by severity of COVID-19, the random effect results demonstrated statistically significant elevation of levels of IL-6 (WMD=18.17 95%CI 3.38 to 32.96, p=0.016), IL-8 (WMD=12.09 95%CI 4.41 to 19.77, p=0.002), MCP-1 (WMD=146.66 95%CI 88.16 to 205.16, p<0.001), CRP (WMD=31.09 95%CI 10.08 to 52.10, p=0.004), PCT (WMD= -31.23 95%CI -37.70 to -24.76, p<0.001), IL-2R (WMD=861.93 95%CI 275.45 to 1448.41, p=0.004), ferritin (WMD= 1083.34 95%CI 431.99 to 1734.70, p=0.001) in severe group as compared to non-severe group. However, the levels of IL-4 (WMD= -18.63 95%CI -51.499 to 14.239, p=0.267), IL-2 (WMD=6.76 95%CI - 0.936 to 14.462, p=0.085), IL-10 (WMD=2.49 95%CI -1.17 to 6.16, p=0.182), TNF-α (WMD= -3.07 95%CI -12.30 to 6.15, p= 0.513), IL-1β (WMD=13.74 95%CI -15.27 to 42.75, p=0.353) were not found significantly associated with the disease severity in COVID-19 patients. Summary effects of all cytokines showed substantial heterogeneity between (I^2^ ∼ 91-97%) contrasting with IL-1β where heterogeneity remained (I^2^ ∼ 62%) in the patient groups as severe and non-severe across all 21 studies **(Table-II)** ^4,4,8,8,17,24–32^.

**Table-II.** Results of meta-analysis comparing laboratory abnormalities in COVID-19 patients with and without severe illness and mortality.

| Laboratory parameter | Severe vs Non-severe |  |  |  |  | Survivors vs Non- survivors |  |  |  |  |
| --- | --- | --- | --- | --- | --- | --- | --- | --- | --- | --- |
|  | *No. of Studies (patients)* | WMD | 95% CI | *p - value* | *I<sup>2</sup> (%)* | *No. of Studies (patients)* | WMD | 95% CI | *P - value* | *I<sup>2</sup> (%)* |
| IL-6(pg/mL) | 9 (1344) | 18.172 | 3.384 to 32.96 | 0.016 | 93.1 | 2 (465) | -37.142 | -101.654 to 27.37 | 0.259 | 98.9 |
| IL-8(pg/mL) | 5 (961) | 12.094 | 4.411 to 19.777 | 0.002 | 95.2 | 1 (274) | -26.633 | -33.031 to -20.236 | p<0.001 | - |
| IL-10 (pg/mL) | 6 (790) | 2.496 | -1.172 to 6.164 | 0.182 | 91.7 | 1 (274) | -7.6 | -8.934 to -6.266 | p<0.001 | - |
| TNF-α pg/mL (<8.1) | 6 (790) | -3.079 | -12.307to 6.15 | 513 | 97.0 | 1 (274) | -4.6 | -5.715 to -3.485 | p<0.001 | - |
| IL-1β (pg/mL) | 4 (635) | 13.742 | -15.27 to 42.754 | 0.105 | 62.0 | 1 (274) | 22.667 | 8.136 to 37.197 | 0.002 | - |
| MCP-1 (pg/mL) | 1 (69) | 146.67 | 88.166 to 205.17 | 0.535 | - | - | - | - | 0 | - |
| CRP (mg/L) | 13 (3123) | 31.091 | 10.081to 52.101 | 0.004 | 97.6 | 2 (484) | -96.408 | -117.84 to -74.975 | p<0.001 | 70.4 |
| PCT (ng/mL) | 14 (2919) | -31.24 | -37.706 to -24.77 | p<0.001 | 99.3 | 2 (675) | 2.952 | -6.237 to 12.141 | 0.529 | 65.5 |
| IL-2R (U/mL) | 4 (628) | 861.931 | 275.452 to 1448.410 | 0.004 | 99.7 | 1 (274) | -666.067 | -782.543 to -549.591 | p<0.001 | - |
| Ferritin, μg/L (30- | 4 (702) | 1083.3 | 432 to 1734.7 | p<0.001 | 95.0 | 2 (465) | -937.606 | -1084.15 to -791.065 | p<0.001 | 0.0 |
| IL-4 (pg/mL) | 2 (220) | -18.63 | -51.499 to14.239 | 0.267 | 91.2 | - | - | - | 0 | - |
| IL-2 (pg/mL) | 1 (93) | 6.763 | -0.936 to 14.462 | 0.085 | - | - | - | - | 0 | - |

IL-1β -interleukin 1 $\beta$ ; IL-2-interleukin 2; IL-2R-interleukin 2R; IL-4-interleukin 4; IL-6- interleukin 6; IL-8- interleukin 8; IL-10- interleukin 10; CRP- C-reactive protein; TNFα - Tumor Necrosis Factor; PCT-Procalcitonin; MCP-1 Monocyte chemotactic protein1 WMD weight mean difference; CI confidence interval.

### 3.2 Association of Cytokines with mortality

For the patients stratified by survival of COVID-19, the random effect results demonstrated statistically significant elevation of levels of IL-2R (WMD= -666.06 95%CI -782.54 to -549.59, p<0.001), IL-8 (WMD= -26.63 95%CI - 33.031 to -20.236, p<0.001), IL-10 (WMD= -7.60 95%CI -8.93 to -6.26, p<0.001), TNF-α (WMD= -4.60 95%CI - 5.71 to -3.48, p<0.001), IL-1β (WMD=22.66 95%CI 8.13 to 37.19, p=0.002), CRP (WMD= -96.40 95%CI -117.84 to -74.97, p<0.001), and ferritin (WMD= -937.60 95%CI -1084.15 to -791.065, p<0.001) in the survivor group as compared to non-survivor group. However, our observation reported levels of IL-6 (p = 0.259), PCT (p = 0.529) were not significantly associated with the mortality. Substantial heterogeneity was observed in IL-6 (I^2^ ∼ 98.9%) in the patient groups as survivors and non-survivors across 3 studies ^17,26,33^ **(Table-II)**.

### 3.3 Comorbidities

None of the comorbidities were statistically associated with the mortality in COVID-19 patients [hypertension (OR=0.58, 95%CI 0.17 to 1.96), diabetes (OR=0.63, 95%CI 0.20 to 1.93), CVD (OR=0.96, 95%CI 0.07 to 12.57), CHD (OR=2.72, 95%CI 0.59 to 12.50), COPD (OR=0.62, 95%CI 0.15 to 2.53), CLD (OR=0.45, 95%CI 0.13 to 1.50), CKD (OR=0.73, 95%CI 0.01 to 33.19), and cancer (OR=2.22, 95%CI 0.20 to 23.82)] **(Table-III)**.

**Table-III:** Results of meta-analysis comparing co-morbidities, complications, and clinical outcome in COVID-19 patients with and without severe illness and mortality.

| **Comorbidity** | **No of studies (participants)** | **OR** | **95% CI** | **p-value** | **I<sup>2</sup> (%)** |
| --- | --- | --- | --- | --- | --- |
| CHD | 1 (191) | 2.72 | 0.59 to 12.50 | 0.197 | - |
| CKD | 2 (484) | 0.73 | 0.01 to 33.19 | 0.897 | 89.9 |
| CLD | 2 (401) | 0.45 | 0.13 to 1.50 | 0.198 | 9.4 |
| COPD | 2 (401) | 0.62 | 0.15 to 2.53 | 0.507 | 0.0 |
| CVD | 2 (484) | 0.96 | 0.07 to 12.57 | 0.979 | 94.6 |
| Cancer/Tumor | 3 (675) | 2.22 | 0.20 to 23.82 | 0.509 | 77.8 |
| Diabetes | 3 (675) | 0.63 | 0.20 to 1.93 | 0.423 | 85.2 |
| Hypertension | 3 (675) | 0.58 | 0.17 to 1.96 | 0.387 | 90.7 |

CVD-Cardio vascular disease; CHD-Coronary heart disease; COPD-chronic obstructive pulmonary disease; CRD-chronic renal disease; CKD- chronic kidney disease; CLD-chronic liver disease; TB-tuberculosis; OR odds ratio; CI confidence interval

## DISCUSSION

COVID-19 a speedily increasing pandemic declared as an emergency by WHO increasing the burden on medical facilities ^3^ which is usually caused by cytokine storm which is the main factor driving severe clinical course of SARS-CoV-2 with a specific immune response that causes severe pathogenic mechanisms through inflammation in a way of ‘attacking’ the body which leads to severe pneumonia, pulmonary oedema, ARDS, or multiple organ failure, ICU admissions and even death^5,7^. Studies have demonstrated the levels of cytokines can also be related to severity of COVID-19 infection ^5,7,11^. Reports suggest that in patients with severe COVID-19, higher levels of IL-2, IL-10, TNF-α are present in ICU patients compared to non-ICU ^12,14^. Cytokines are quantitative measurements used clinically for many conditions reflecting pathological development of COVID-19 and evidence from literature depicts a major role of interleukins as an useful approach to clinicians for establishing a treatment and close monitoring and improving prognosis and outcomes and their significant variability between different groups of patients ^5^. Cytokines drive T-cell growth, induces the differentiation of regulatory T-cells, and mediates activation-induced cell death ^34,35^. IL-4 (IL-4R) signaling plays an essential role in immune responses through their signaling via type I and type II IL-4Rs on neutrophils to inhibition of several neutrophil effector functions fighting against infection at the site of infection^36^. IL-8 can add to neutrophil-mediated tissue damage, inflammatory diseases and host defense, neutrophils, granulocytes, causing migration toward the site of infection. ^37^. IL-10, which is also to referred as human cytokine synthesis inhibitory factor (CSIF), acts as an anti-inflammatory cytokine ^38^. IL-6 is an interleukin that acts as a pro-inflammatory cytokine^15,39^. Elevated levels of IL-6 can cause hyper-activation of JAK/STAT3 signaling, which is often associated with poor patient outcomes ^16,40,41^, implying a possible shared mechanism of cytokine-mediated lung damage caused by COVID-9 infection ^42,43^ along with co-morbidities like hypertension in severe group of patients more than non-severe ones is contributing to disease severity and case fatality ^32^. Therefore, it is imperative to recognize the markers observing the progression of disease and treat patients early. The elevated cytokine levels may also be responsible for the lethal complications of patients with COVID-19 ^17^. Severity in COVID-19 may also be associated with IFN-γ production by CD4+ T-cells as its defect may lead to hypercytokinaemia ^12^. Along with interleukins, the levels of C-reactive protein (CRP), induced by IL-6, is a significant biomarker of inflammation, infection and tissue damage ^18^. The correlation between CRP levels, lung lesions, and disease severity can provide reference for clinical treatment ^18,40^. Procalcitonin (PCT) that is a glycoprotein and is the precursor of calcitonin, its levels are increased with bacterial and viral levels and can be indicative of the disease severity as these levels were found to be greater in severe group of patients than in mild group ^9,32^indicating its association with disease severity and mortality^4,6,8,19,24,27,28,30,32,33,44–47^. Ferritin, a protein that contains iron and is the primary form of iron stored inside of cells. Levels of ferritin could be a significant marker for COVID-19 as it is found elevated in diseased patients^19^(S. Li et al., 2020;, Wu, Ke et al., 2020)^4,30,32,48^. Nevertheless, the role of inflammatory markers in observing the severity and mortality of COVID-19 is still controversial. In this study, through examining the 21 retrospective studies, we concluded that inflammatory markers were positively correlated with the severity and mortality of COVID-19.

Furthermore, categorizing patients into severe and non-severe sub-groups is dependent on the definitions of severity of COVID-19. The approach was to measure severity of the disease between two groups as it was found from different literature studies. A retrospective study of 552 patients stated panel of circulating cytokinesIL-2R, IL-6, IL-8, IL-10, IL-1β and TNFα, PCT, CRP and ferritin could predict disease deterioration as the levels of cytokines were greatly increased in the severe group of patients than the non-severe ones which in turn was associated with the severity and mortality risk ^46^. A meta-analysis demonstrated IL-6, IL-10 and serum ferritin were significantly elevated in patients with both severe and fatal COVID-19 indicating these interleukins as a strong discriminators for severe disease^49,50^. Increased CRP, IL-6 and TNF-α in COVID-19 patients and has shown a significant role of these markers in disease prognosis ^31^. Furthermore, a retrospective study on 83 patients demonstrated how cytokines can be a targeted approach for the treatment methods for COVID-19 with IL blocking agents for severely ill patients seems effective as it is essential to accomplish a treatment for patient’s condition in a timely manner^51^. Moreover, strategies that encourage pulmonary recruitment and over activation of inflammatory cells by suppressing cytokine storm might improve the outcomes of severe COVID-19 patients ^8^. The above discussed findings show that more attention is warranted when interpreting cytokine findings in patients with COVID-19 as patients with highly increased levels require proper management. Furthermore, studies based on patients with co-morbidities like diabetes and hypertension, and age above 50 demonstrated greater risk of death in COVID-19 patients^52^.

Multiple studies on COVID-19 have concurred high amount of information on cytokines and their use on the basis of their role in severity of the disease which independently has been suggested to give a potential approach for treatment of COVID-19 as patients have consistently high levels of cytokines, with significant co-morbidities such as diabetes, hypertension and CVD reported in critically ill patients. Therefore, further research on the same can give a better insight to the clinicians regarding the disease progression of patients. However, the role of inflammatory markers in monitoring the severity of COVID-19 is still controversial ^12^.

A copious amount of data was collected from different locations where different levels were seen in different patients amongst the same set of parameters taking in consideration the variability the study. Variability can be accounted to the differences in patient exposure to the virus. To ascertain the usefulness of the cytokines listed in this systematic review as indicators of disease progression, and whether they definitively rise in COVID-19 may require further data collection.

There are several limitations that needs to be mentioned. The number of studies included in the meta-analysis is small. There was heterogeneity amongst individual studies because of which there was a deviation of some of our results from usual findings. Additionally, case-series were included in the present meta-analysis. Although we did an extensive search, we may have inadvertently missed relevant studies. Exclusion of studies in languages other than English may have resulted in missing of relevant studies.

## CONCLUSION

The present systematic review will provide understanding of the role of inflammatory markers in the pathophysiology of the disease COVID-19 which is associated with age and comorbidities donating to disease severity and mortality. The uncovering of these clinical parameters can contribute in the suitable diagnostic and treatment approach in a timely manner that can further help in the reduction of the mortality rate in the patients of COVID-19. Also, most of the studies referenced in this systematic review are cohort studies originating in Wuhan, China, but because the virus is now global pandemic it thus requires international studies with a bigger sample size with role of inflammatory markers in the disease diagnosis and prognosis.

## Data Availability

The author confirms that the data supporting the findings of the study are available within the article [and/ or its supplementary material].

## DECLARATIONS

### Funding

This research did not receive any specific grant from funding agencies in the public, commercial, or not-for-profit sectors.

### Conflicts of interest/Competing interests

None

### Code availability (software application or custom code)

Not applicable

### Authors’ contributions

Conception and design-**Nidhi and Pinki**; acquisition of data-**Sara and Pinki**; analysis and interpretation of data-**Rizwana and Ram**; drafting the article-**Sara and Pinki**; revised article critically for important intellectual content-**Nidhi and Rizwana**; final approval of the version to be published-**Nidhi**

## ICMJE DISCLOSURE FORM

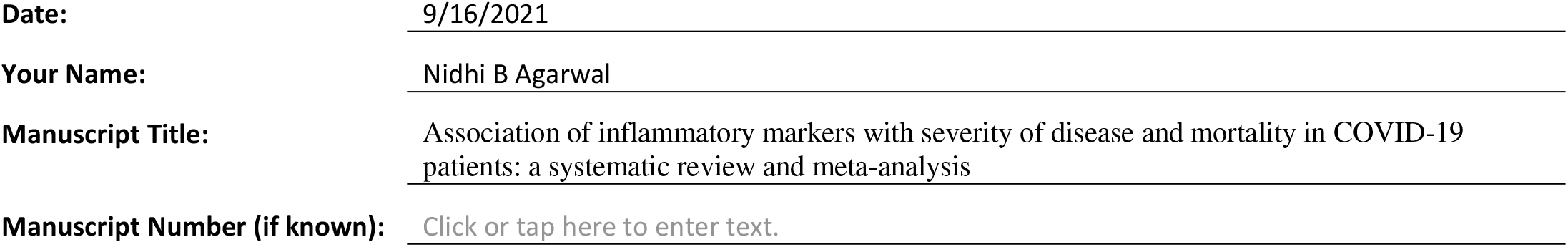

In the interest of transparency, we ask you to disclose all relationships/activities/interests listed below that are related to the content of your manuscript. “Related” means any relation with for-profit or not-for-profit third parties whose interests may be affected by the content of the manuscript. Disclosure represents a commitment to transparency and does not necessarily indicate a bias. If you are in doubt about whether to list a relationship/activity/interest, it is preferable that you do so.

In the interest of transparency, we ask you to disclose all relationships/activities/interests listed below that are related to the content of your manuscript. “Related” means any relation with for-profit or not-for-profit third parties whose interests may be affected by the content of the manuscript. Disclosure represents a commitment to transparency and does not necessarily indicate a bias. If you are in doubt about whether to list a relationship/activity/interest, it is preferable that you do so.

The author’s relationships/activities/interests should be defined broadly. For example, if your manuscript pertains to the epidemiology of hypertension, you should declare all relationships with manufacturers of antihypertensive medication, even if that medication is not mentioned in the manuscript.

In item #1 below, report all support for the work reported in this manuscript without time limit. For all other items, the time frame for disclosure is the past 36 months.

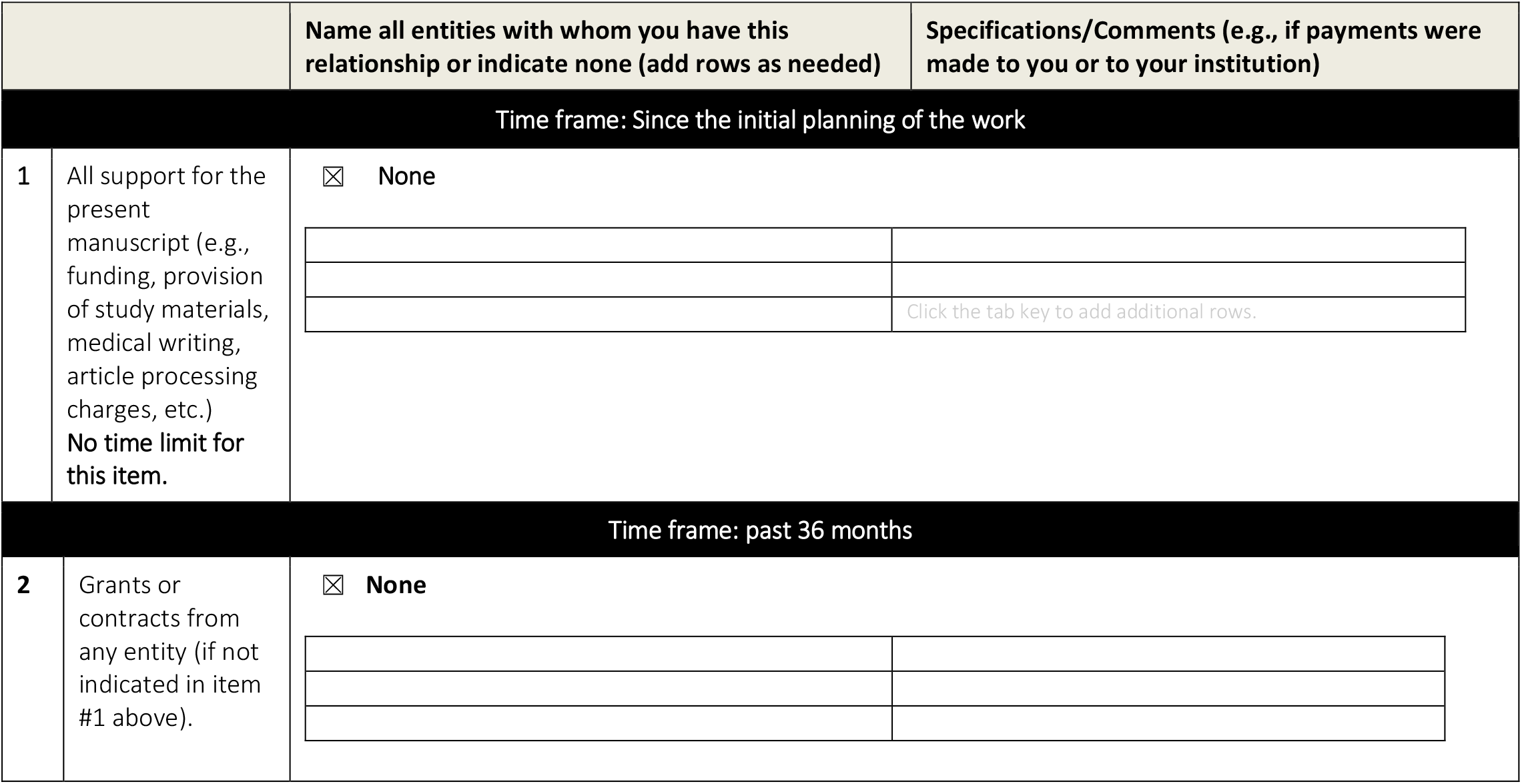
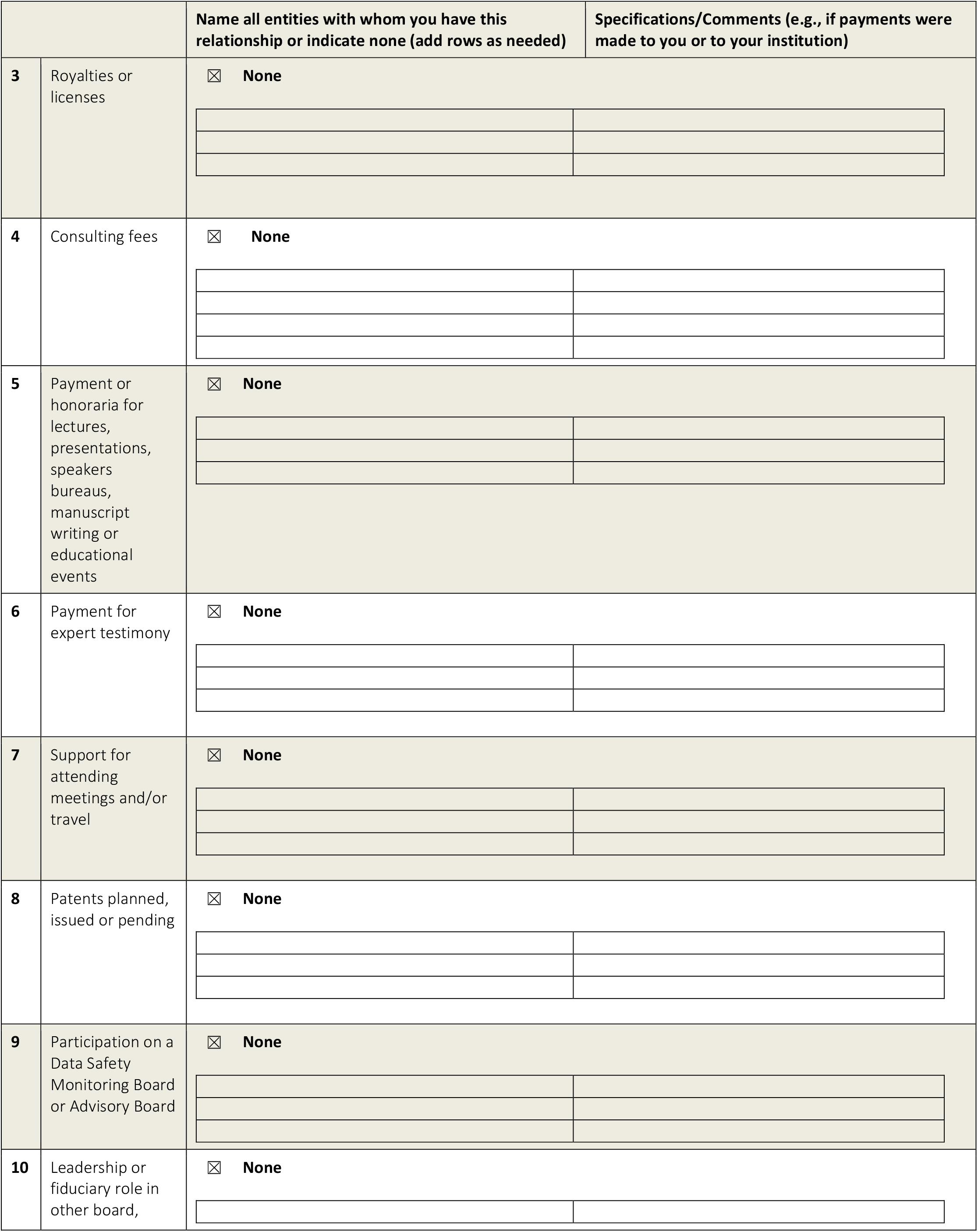
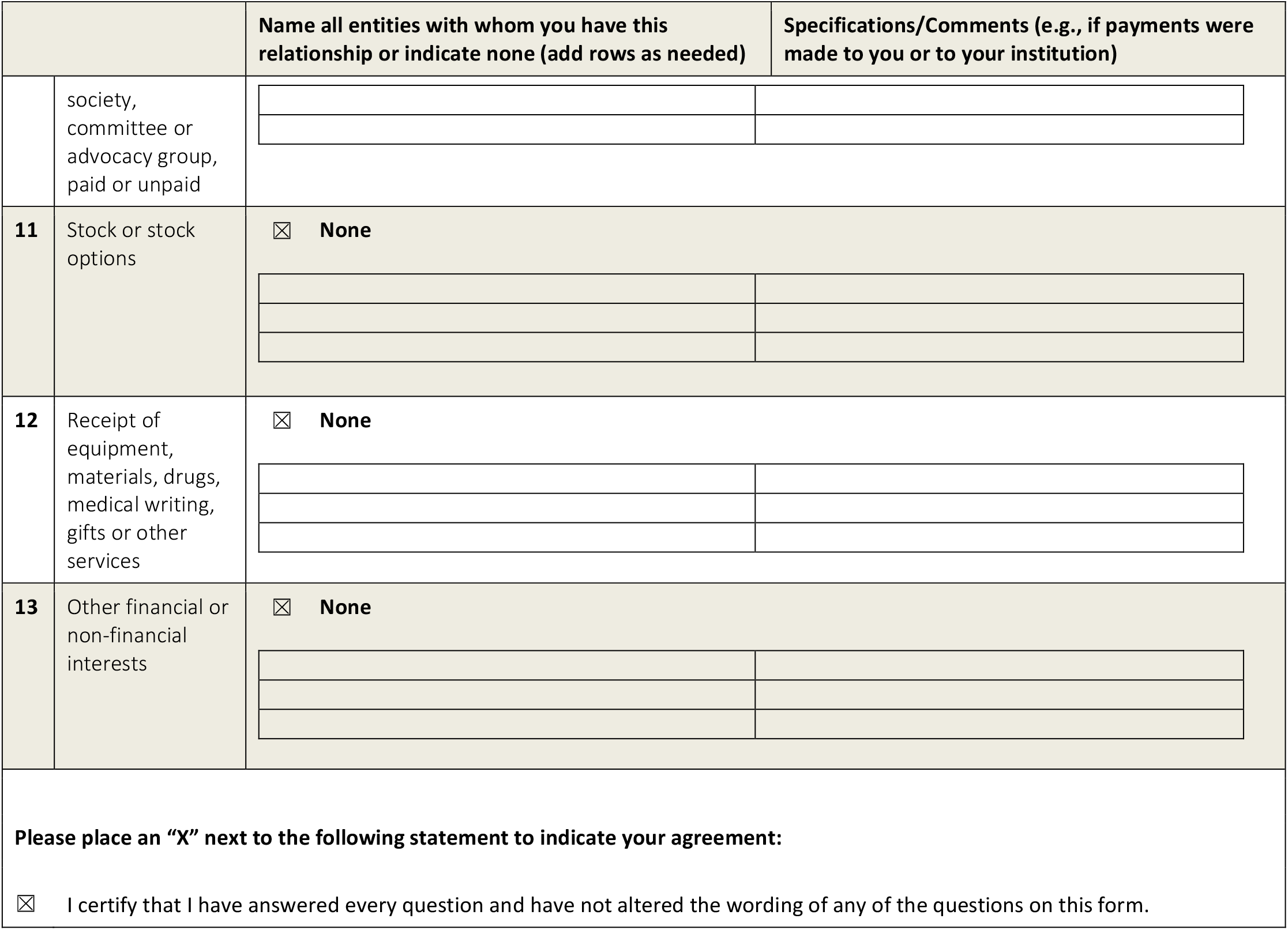

### Preferred Reporting Items for Systematic reviews and Meta-Analyses extension for Scoping Reviews (PRISMA-ScR) Checklist

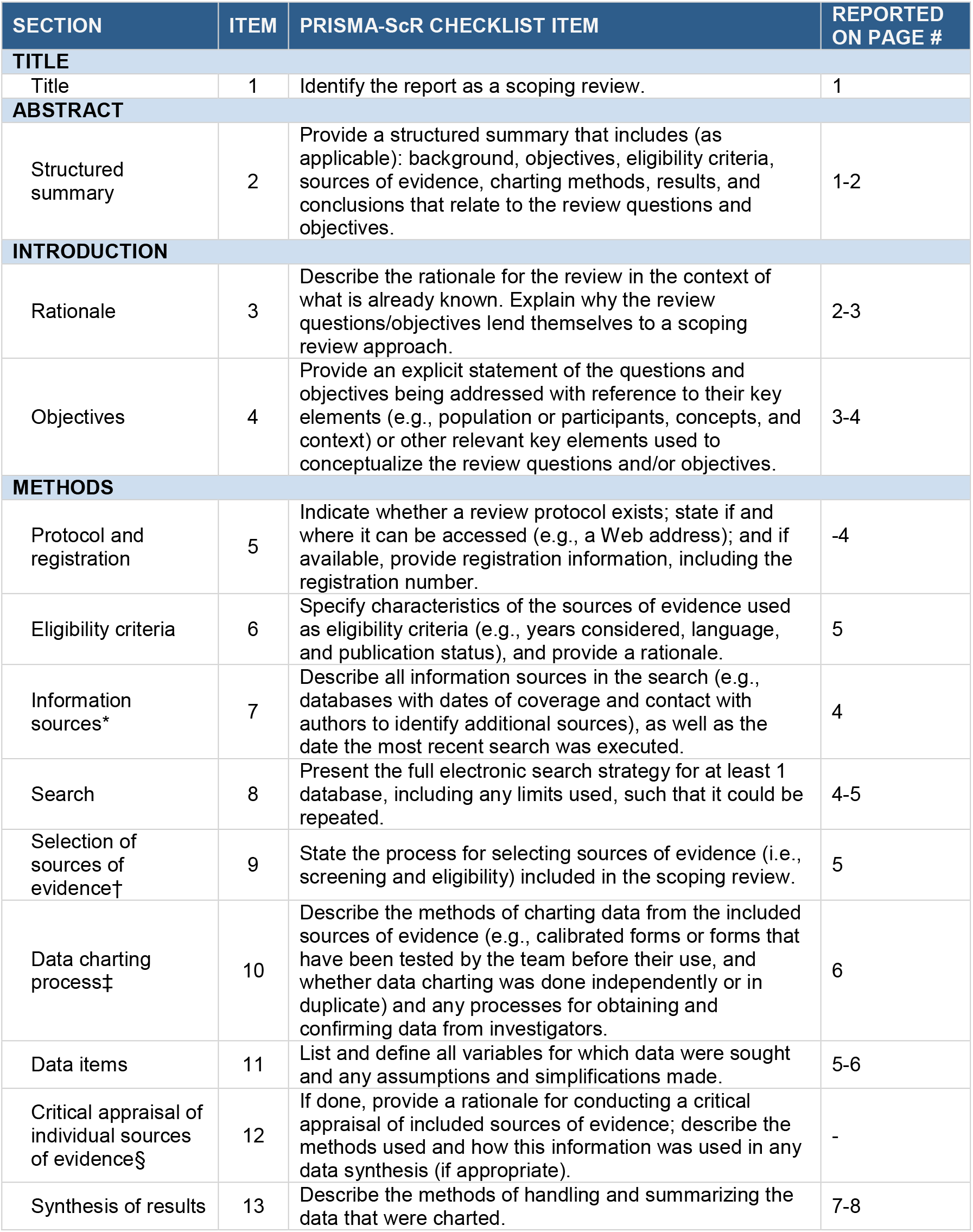
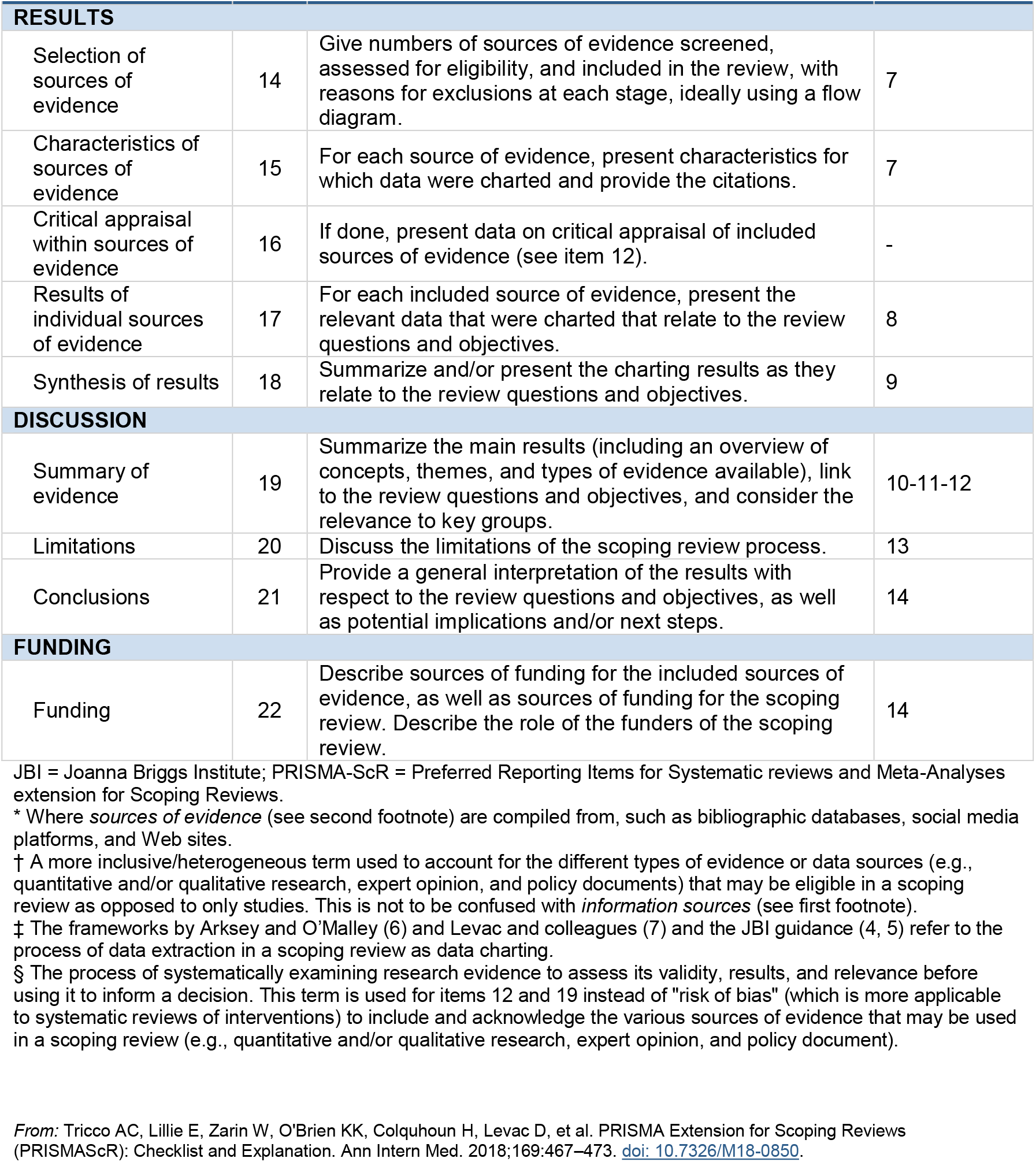

## Notes

### Competing Interest Statement

The authors have declared no competing interest.

