## Supplementary material for "Association of inflammatory markers with severity of disease and mortality in COVID-19 patients: a systematic review and meta-analysis"

Jamia Hamdard

New Delhi-110062, India

**1. Quality assessment of Included Studies Using NIH tool of quality assessment**

*Quality assessment of cohort and cross-sectional studies*

| **Criteria** | **Cai**  **et al 2020** | **Qin et al 2020** | **Wu et al 2020** | **Li et al 2020** | **Zhang**  **et al 2020** | **Huang**  **et al 2020** | **Gao et al 2020** | **Chen**  **et al 2020** | **Tao**  **et al 2020** | **Gao**  **et al 2020** | **Guan**  **et al 2020** | **Yang**  **et al 2020** | **Zhou**  **et al 2020** | **Li et**  **al 2020** | **Chen**  **et al 2020** | **Herold**  **et al 2020** |
| --- | --- | --- | --- | --- | --- | --- | --- | --- | --- | --- | --- | --- | --- | --- | --- | --- |
| Was the research question or objective in this paper clearly  stated? | Yes | Yes | Yes | Yes | Yes | Yes | Yes | Yes | Yes | Yes | Yes | Yes | Yes | Yes | Yes | Yes |
| Was the study population clearly specified and defined? | Yes | Yes | Yes | Yes | Yes | Yes | Yes | Yes | Yes | Yes | Yes | Yes | Yes | Yes | Yes | Yes |
| Was the participation rate of eligible persons at  least 50%? | NA | Yes | Yes | Yes | Yes | Yes | Yes | No | Yes | Yes | Yes | Yes | No | Yes | Yes | Yes |
| Were all the subjects selected or recruited from the same or similar populations (including the same time period)? Were inclusion and exclusion criteria for being in the study prespecified  and applied | Yes | Yes | Yes | Yes | Yes | Yes | Yes | Yes | Yes | Yes | Yes | Yes | Yes | Yes | Yes | Yes |

| uniformly to all  participants? |  |  |  |  |  |  |  |  |  |  |  |  |  |  |  |  |
| --- | --- | --- | --- | --- | --- | --- | --- | --- | --- | --- | --- | --- | --- | --- | --- | --- |
| Was a sample size justification, power description, or variance and effect  estimates provided? | NA | NA | NA | NA | NA | NA | NA | NA | NA | NA | NA | NA | NA | NA | NA | NA |
| For the analyses in this paper, were the exposure(s) of interest measured prior to the outcome(s) being  measured? | No | Yes | Yes | Yes | Yes | Yes | Yes | No | Yes | Yes | Yes | Yes | No | Yes | Yes | Yes |
| Was the timeframe sufficient so that one could reasonably expect to see an association between exposure and outcome if it  existed? | Yes | Yes | Yes | Yes | Yes | Yes | Yes | Yes | Yes | Yes | Yes | Yes | Yes | Yes | Yes | Yes |
| For exposures that can vary in amount or level, did the study examine different levels of the exposure as related to the outcome (e.g., categories of exposure, or  exposure measured | NA | NA | NA | NA | NA | NA | NA | NA | NA | NA | NA | NA | NA | NA | NA | NA |

| as continuous  variable)? |  |  |  |  |  |  |  |  |  |  |  |  |  |  |  |  |
| --- | --- | --- | --- | --- | --- | --- | --- | --- | --- | --- | --- | --- | --- | --- | --- | --- |
| Were the exposure measures (independent variables) clearly defined, valid, reliable, and implemented consistently across all study  participants? | Yes | Yes | Yes | Yes | Yes | Yes | Yes | Yes | Yes | Yes | Yes | Yes | No | Yes | Yes | Yes |
| Was the exposure(s) assessed more than once over time? | NA | NA | NA | NA | NA | NA | NA | NA | NA | NA | NA | NA | NA | NA | NA | NA |
| Were the outcome measures (dependent variables) clearly defined, valid, reliable, and implemented consistently across all study participants? | Yes | Yes | Yes | Yes | Yes | Yes | Yes | Yes | Yes | Yes | Yes | Yes | No | Yes | Yes | Yes |
| Were the outcome assessors blinded to the exposure status  of participants? | NA | NA | NA | NA | NA | NA | NA | NA | NA | NA | NA | NA | NA | NA | NA | NA |
| Was loss to follow-  up after baseline 20% or less? | NA | NA | NA | NA | NA | NA | NA | NA | NA | NA | NA | NA | NA | NA | NA | NA |

| Were key potential confounding variables measured and adjusted statistically for their impact on the relationship between exposure(s) and  outcome(s)? | No | Yes | Yes | Yes | Yes | Yes | Yes | No | Yes | Yes | Yes | Yes | Yes | Yes | Yes | Yes |
| --- | --- | --- | --- | --- | --- | --- | --- | --- | --- | --- | --- | --- | --- | --- | --- | --- |
| Quality Rating (CD, cannot determine; NA, not applicable;  NR, not reported | Fair | Good | Good | Good | Good | Good | Good | Fair | Good | Good | Good | Good | Fair | Good | Good | Good |

*Quality assessment of case-series*

| **Criteria** | **Shaohua et al 2020** | **Wang et al 2020** | **Zhua et al 2020** | **Wan et al 2020** | **Zhao et al 2020** |
| --- | --- | --- | --- | --- | --- |
| Was the study question or objective clearly  stated? | Yes | Yes | Yes | Yes | Yes |
| Was the study population clearly and fully described, including a case  definition? | Yes | Yes | No | No | Yes |
| Were the cases  consecutive? |  |  |  |  |  |

| Were the subjects  comparable? | Yes | Yes | Yes | Yes | Yes |
| --- | --- | --- | --- | --- | --- |
| Was the intervention  clearly described? | NA | NA | NA | NA | NA |
| Were the outcome measures clearly defined, valid, reliable, and implemented consistently across all  study participants? | Yes | Yes | Yes | No | Yes |
| Was the length of  follow-up adequate? | Yes | Yes | No | Yes | Yes |
| Were the statistical  methods well-described? | Yes | Yes | Yes | Yes | Yes |
| Were the results well-  described? | Yes | Yes | Yes | Yes | Yes |
| Quality Rating (CD, cannot determine; NA, not applicable; NR, not  reported | Good | Good | Fair | Fair | Good |
